# Effects of a 5-week heart rate biofeedback randomized intervention on texture in the Alzheimer’s Disease signature cortical region

**DOI:** 10.64898/2026.06.03.26354853

**Authors:** Seo Yeon Lee, Kaoru Nashiro, Jungwon Min, Hyun Joo Yoo

## Abstract

Using data from a randomized clinical trial, we examined whether daily biofeedback training that modulates heart rate oscillations is associated with changes in microstructural brain texture in Alzheimer’s disease–signature cortical (ADSC) and hippocampal regions. Younger and older adults were randomly assigned to one of two daily biofeedback practices for five weeks: slow-paced breathing designed to increase heart rate oscillations (Osc+) or self-selected strategies aimed at decreasing oscillations (Osc−). Intervention effects were observed in both ADSC and hippocampus regions and were confined to a composite texture factor dominated by uniformity and entropy. Across regions, effects were expressed primarily as Time × Condition interactions, indicating differential texture trajectories between Osc+ and Osc−. In the hippocampus, this pattern was further qualified by a Time × Condition × Age Group interaction, reflecting more pronounced effects in older adults, whereas younger adults showed no reliable texture modulation. Partial least squares correlation analyses further demonstrated that training-related texture changes in the left hippocampus, right fusiform gyrus, and right entorhinal cortex covaried with concurrent changes in plasma AD-related biomarkers, with tau- and p-tau–related measures contributing most strongly to the multivariate association. Together, these findings suggest that HRV biofeedback may selectively influence specific dimensions of brain microstructural texture and that such changes are meaningfully coupled with plasma AD-related biomarker profiles.

## 1. Introduction

With the rise in global population and life expectancy, age-related disorders, especially Alzheimer’s disease (AD), have emerged. More than 50 million individuals in the world are estimated to have AD (Livingston et al., 2020). This growing burden is closely linked to aging, which has been largely considered the primary risk factor for AD (Hou et al., 2019). Consistent with this, the incidence of AD increases 2-to 3-fold with each additional decade of age (Rajan et al., 2021).

With aging, the brain undergoes various structural changes, such as cerebral atrophy, gray and white matter changes, volume loss, ventricular enlargement, and sulci widening (Blinkouskaya & Weickenmeier, 2021; J. Lee & Kim, 2022). Recent studies have demonstrated that the volume of the brain decreases at an increasing rate per decade after 40 years old (DeCarli et al., 2005; J. Lee & Kim, 2022; Peters, 2006). The grooves and folds of the sulci and gyrus, respectively, of the cerebral cortex become wider and shallower, respectively, with aging (Jin et al., 2018). These macroscopic changes, however, are the consequence of abnormal deposition of amyloid plaques and tau pathology rather than a direct measure of AD progression (Wearn et al., 2023). In widely used biological frameworks of AD such as the A/T/N (amyloid/tau/neurodegeneration) model, amyloid-β deposition (A) and tau pathology (T) occur earlier in the disease cascade, whereas neurodegeneration (N), including structural atrophy detectable by MRI, is considered a downstream biomarker that becomes abnormal later in the pathogenic sequence (Jack et al., 2016; Jack Jr. et al., 2024). In other words, amyloid and tau abnormalities precede marked volume loss. This ordering implies that detectable changes in brain volume emerge at a state that is already irreversible during the pathogenic progression of aging and AD (Jack et al., 2016; Jack Jr. et al., 2024). Consequently, volumetric measures may lack sensitivity to early alterations associated with amyloid and tau pathology, underscoring the need for imaging earlier microstructural changes.

An analytic approach that may signal relevant and timely structural alterations in AD is MRI texture analysis (Cai et al., 2020a). This approach provides a quantitative approach to characterize tissue properties. Specifically, subtle changes in the MR signal, including smoothness, randomness, and heterogeneity, can be quantified using texture analysis (Cai et al., 2020a; Wearn et al., 2023). Several studies have shown that texture analysis can predict the future conversion of people with mild cognitive impairment (MCI) to a diagnosis of AD (Gao et al., 2018; Luk et al., 2018; Sørensen et al., 2015). Specifically, hippocampal texture individually, much like shape and volume, was shown to be a significant predictor of development of dementia (Achterberg et al., 2019; Wearn et al., 2023). Moreover, Sørensen et al. found a significant correlation between texture and hippocampal FDG-PET uptake, or brain metabolism, and hippocampal MRI texture remained significant after decorrelating volume. In other words, texture is an independent marker beyond hippocampal shrinkage, which may capture pathological changes not easily reflected by volumetric atrophy (Sørensen et al., 2015). A direct analysis of texture from MRI, therefore, has the potential to detect subtle brain changes linked with AD pathology before clinical diagnosis of dementia.

Accordingly, biomarkers potentially underlying these textural alterations are plasma and cerebrospinal fluid (CSF) amyloid beta (Aβ) and phosphorylated tau proteins, which have been strongly associated with AD pathology and essential in screening, monitoring, and serving as therapeutic targets in AD (Jangampalli Adi Pradeepkiran, 2025; Pais et al., 2023). Plasma and CSF Aβ levels are significantly altered in AD patients, suggesting the potential of these biomarkers to contribute to diagnosis (Jangampalli Adi Pradeepkiran, 2025; Pais et al., 2023; Smirnov et al., 2022). Specifically, mutations in the amyloid precursor protein and presenilin resulted in buildup of Aβ plaques, while apolipoprotein E4 impaired Aβ clearance (Hardy & Selkoe, 2002; Jangampalli Adi Pradeepkiran, 2025; Selkoe & Hardy, 2016). Together, they form an imbalance of production and clearance of Aβ, worsening AD progression, which also alters patterns observable in neuroimaging (Hardy & Selkoe, 2002; Jangampalli Adi Pradeepkiran, 2025; Leandrou et al., 2020; Selkoe & Hardy, 2016). Therefore, subtle changes, i.e. texture alterations related to deposition of AB peptide and neurofibrillary tangles before overt brain atrophy occurs, may be quantified using texture analysis of AD signature cortical (ADSC) regions (Cai et al., 2020b; Leandrou et al., 2020; Wearn et al., 2023). This early detection through texture and Aβ and tau pathology offers the possibility for timely intervention and improved disease management.

Our study focuses on the hippocampal and ADSC regions, which include the bilateral inferior and middle temporal lobes, entorhinal cortex and fusiform gyrus, which are recognized as key regions of interest (ROIs) for neuroimaging analyses because of their early and substantial involvement in AD (Dickerson et al., 2009; Schwarz et al., 2016). Measurements of cortical thinning, volume changes, and texture features within these ROIs correlate with symptom severity and AD progression.

Importantly, accumulating evidence suggests that these structural characteristics are not only disease markers but may also be influenced by physiological regulation. In particular, individual differences in resting heart rate variability (HRV) have been linked to cortical integrity, raising the possibility that HRV interventions could modulate ADSC features (Carnevali et al., 2019; Koenig et al., 2021; Wood et al., 2017; Yoo et al., 2018). Datasets from our lab indicate that greater structural thickness in OFC regions was associated with greater HRV in young and older adults (Koenig et al., 2018; Yoo et al., 2018). In addition, in a recent clinical trial, we found that 5 weeks of HRV biofeedback training increased left amygdala-medial PFC connectivity and generally increased functional connectivity within emotion-related resting-state networks in younger adults (ClinicalTrials.gov NCT03458910; Heart Rate Variability and Emotion Regulation or “HRV-ER”; Nashiro et al., 2022). Consistent with prior correlations between individual differences in left orbitofrontal cortex structure and HRV (Koenig et al., 2021; Yoo et al., 2018), the interventions differentially affected structural volume in left orbitofrontal cortex regions in both the younger and older adult cohorts (Yoo et al., 2022). In older adults, the interventions differentially affected volume in LC-innervated-regions hippocampal ROI (i.e., the molecular layer, CA3, CA4 and granule cell layer of dentate gyrus; Yoo et al., 2023).

In the current study, we followed up on these prior findings to examine whether HRV interventions influence texture features in ADSC and hippocampal regions. Participants were assigned to conditions designed to increase or decrease heart rate oscillations, allowing us to test whether daily HRV biofeedback training led to measurable changes in MRI-derived texture. We took a region-of-interest (ROI) approach selecting bilateral ADSC (entorhinal, inferior temporal, middle temporal, and fusiform cortices) and hippocampal regions (Dickerson et al., 2009; Schwarz et al., 2016). Additionally, we examined the age differences in HRV training effects on texture in ADSC and hippocampal regions. Finally, we investigated multimodal coupling between plasma biomarkers and structural changes, testing whether reductions in Aβ and tau species were associated with alterations in ADSC and hippocampal texture. Based on these aims, we hypothesized that an Osc+ condition in the HRV biofeedback training would induce enhanced texture integrity within ADSC and hippocampal regions due to associations with better autonomic regulation and cerebral blood flow. We also expect to see that age would moderate these effects, where older adults would demonstrate larger oscillation-related changes in texture than older adults. Thereby, improvements in hippocampal and ADSC texture in the Osc+ condition would be associated with reductions in plasma Aβ and tau biomarkers.

## 2. Methods

### 2.1. Participants

The current study was part of a larger heart rate variability biofeedback intervention study (ClinicalTrials.gov NCT03458910; Heart Rate Variability and Emotion Regulation or “HRV-ER”; (Nashiro et al., 2023)). In this clinical trial, we recruited 121 younger participants (aged 18-35 years) and 72 older participants (aged 55-80 years) via the USC Healthy Minds community subject pool, a USC online bulletin board, Facebook and flyers between January 2018 and March 2020; the current study focused on the 52 younger participants and 48 older participants who had blood samples and MRI scans at both pre and post intervention.

Participants provided written informed consent approved by the University of Southern California (USC) Institutional Review Board. Participants were assigned to small groups of 3-6 people, with each group meeting at the same time and day each week. After recruitment and scheduling of each wave of groups, the groups were randomized to one of two conditions (see Supplementary Figure S1 for flow diagram): biofeedback training either to increase or decrease HRV. Upon completing the study, participants were paid for their participation and received bonus payments based on their individual and group performances (incentives for training were the same across conditions; see details below under “Rewards for Performance”). Prospective participants were screened and excluded for any medical, neurological, or psychiatric illness (but we included people who were taking antidepressant or anti-anxiety medication and/or attending psychotherapy only if the treatment had been ongoing and unchanged for at least three months and no changes were anticipated). We excluded people who had a disorder that would impede performing the HRV biofeedback procedures (e.g., coronary artery disease, angina, cardiac pacemaker), who were currently trained in relaxation, biofeedback or breathing techniques, or were on any psychoactive drugs other than antidepressants or anti-anxiety medications. For older adults, we also excluded people who scored lower than 16 on the TELE (Gatz et al., 1995) for possible dementia. Gender, education, age, and race were similar in the two conditions.

### 2.2. Procedures

#### 2.2.1. Overview of 7-week Protocol Schedule

The study protocol involved seven weekly lab visits and five weeks of home biofeedback training. The first lab visit assessed the non-MRI baseline measurements, including various questionnaires. The second lab visit involved the baseline MRI session with the first biofeedback calibration and training session. Each of the lab calibration sessions began with a 5-min baseline rest period followed by different strategies to find the optimal condition. After calibration sessions were completed, participants were informed of the best strategy and instructed to practice the best condition at home twice daily. Practice duration was 10 minutes twice a day for the first training week (between the 1st-week visit and the 2nd-week visit), 15 minutes twice a day for the second training week (between the 2nd week visit and the 3rd week visit), and 20 minutes twice a day for the last weeks (between the 3rd week visit and the 7th week visit).

The weekly lab visits (except for weeks with MRI sessions) were conducted in small groups consisting of participants assigned to the same condition. During these sessions, participants shared their experiences and strategies about biofeedback training, facilitated by 1-2 researchers. Outside the lab, participants used a customized social app to communicate with other members of their group and researchers about their progress on daily biofeedback training. For instance, participants exchanged positive feedback such as ‘thumbs-up’ or smiling face emojis upon completing training for the day, while researchers provided friendly reminders when participants were falling behind in completing their home practice. The week-6 lab visit repeated the assessments from the initial lab visit. The final (7th) lab visit began with a repeat of the baseline MRI session scans in the same order.

#### 2.2.2. Biofeedback Training for the Osc+ condition

During all practice sessions, participants wore an ear sensor to measure their pulse. They received real-time heart rate biofeedback while breathing in through the nose and out through the mouth, synchronized with the emWave pacer. The emWave software (HeartMath®Institute, 2020) generated a summary ‘coherence’ score defined as peak power/(total power - peak power). Peak power was derived by finding the highest peak within the range of .04 - .26 Hz and calculating the integral of the window .015 Hz above and below this highest peak, divided by total power computed for the .0033 - .4 Hz range.

During the second lab visit, we introduced participants to the device, and individual resonance frequency was identified during five minutes of paced breathing at 6, 6.5, 5.5, 5 and finally 4.5 breaths/min (Lehrer et al., 2013). Following completion of all 5-min breathing segments, we computed various aspects of the oscillatory dynamics for each breathing pace using Kubios HRV Premium 3.1 software (Tarvainen et al., 2014). Then, we estimated which breathing pace most strongly approximated the resonance frequency by assessing which one had the most of the following characteristics: highest low frequency (LF) power, the highest maximum LF amplitude peak on the spectral graph, highest peak-to-trough amplitude, cleanest and highest-amplitude LF peak, highest coherence score and highest the root mean squared successive differences (RMSSD). Participants were then instructed to practice at home using the pacer set to their identified individual resonance frequency and to attempt to maximize their coherence scores.

During the third visit, participants completed three 5-min paced breathing segments: the best condition from the last week’s visit, half breath per minute faster and half breath slower than the best condition. They were then instructed to continue home practice at the pace that best approximated the resonance frequency based on the characteristics listed above the following week. In subsequent weekly visits, they were asked to experiment with abdominal breathing and inhaling through nose/exhaling through pursed lips as well as other strategies of their choice during 5-min training segments.

#### 2.2.3. Biofeedback Training for the Osc-condition

The same biofeedback ear sensor was used in this condition. However, we created custom software to display an alternative set of feedback to the Osc-participants. During each Osc-training session, a ‘calmness’ score was provided as feedback to the participants instead of the coherence score. The calmness score was calculated by multiplying the coherence score that would have been displayed in the Osc+ condition by −1 adding 10 (an ‘anti-coherence’ score). Thus, participants received more positive feedback (higher calmness scores) when their heart rate oscillatory activity in the 0.04 – 0.26 Hz range was low.

Participants also received a point adjustment that gave a penalty when heart rate was the lowest it had been in the past 15 s. Specifically, every 5 s, a local maximum IBI was set based on the maximum IBI from the last 15 s. If, at that point, the participant’s current IBI was longer than this local maximum, the calmness score displayed for the next 5 s was the anti-coherence score - 2. Naturally, most of the time, current IBI was lower than the local maximum, and in those cases, the calmness score was the anti-coherence score +1. Thus, there was a penalty in their calmness score for periods when their heart rate was slower than it had been in any of the past 15 s. Nevertheless, the average heart rate during biofeedback sessions did not differ significantly across conditions, suggesting that this additional feedback has had little impact on heart rate.

During the initial calibration session at the end of the second lab visit, participants were introduced to the device and feedback and were asked to come up with five strategies aimed at lowering heart rate and heart rate oscillations. Each participant was instructed to wear the ear sensor and view real-time heart rate biofeedback while they tried each strategy for five minutes. We analyzed the data in Kubios and identified the best strategy as the one that had the most of the following characteristics: lowest LF power, the minimum LF amplitude peak on the spectral graph, lowest peak-to-trough amplitude, multiple and lowest-amplitude LF peak, highest calmness score and lowest RMSSD. Participants were then instructed to use this strategy to maximize their calmness scores in their home training sessions.

On the third visit, they were instructed to select three strategies and test each out in a 5-min session. The strategy identified as the best (based on the same characteristics used in the initial calibration session) was selected as the one to focus on during home training. In subsequent weekly visits, during 5-min training segments, they were asked to try out strategies of their choice.

#### 2.2.4. Blood Collection Procedure

The HRV biofeedback intervention lasted for 7 weeks. The baseline measurements were collected during the first and second week, the intervention technique was introduced to each participant during the second week, and the post-intervention measures were collected during the sixth and seventh week. The blood samples were collected via antecubital venipuncture during the first- and sixth-week sessions. A professional phlebotomist drew 10 ml of blood into a K2 EDTA tube from each participant’s arm and 2.5 ml of blood into a PAXgene RNA tube. The whole blood from the K2 EDTA tubes was then centrifuged at the speed of 1500 RPM for 15 minutes at room temperature (15°C) to separate plasma from blood cells. Plasma was then allocated in cryovials and stored at −80°C until transferred and assayed at University of California, Irvine for Aβ42 and Aβ40. The PAXgene RNA tubes were gently inverted following collection and kept at room temperature for between 2.00 and 70.2 hours (mean = 6.95 hours; 86% of the samples were within 2-3 hours) before storing them at −80°C.

#### 2.2.5. Rewards for Performance

In addition to receiving compensation of $15 per hour for each lab visit, participants were eligible for performance-based incentives at both the individual and group levels. For individual rewards, participants could earn $2 each week for each time (up to a maximum of 10) they exceeded their assigned target score (target scores were assigned each week and were the average of the top 10 scores earned from the previous week’s training sessions plus 0.3). Group performance rewards were earned when members of a participant’s group completed a minimum of 80% of their assigned biofeedback training minutes. For example, if a participant completed 100% of their training, they received an additional $3 for each group member who also completed 100% of their training. If a participant completed 80% of their training, they received an additional $2 for each group member who also completed at least 80% of their training. Rewards were calculated weekly, and participants received weekly updates on their earnings at their lab visits.

### 2.3. MRI Scan Parameters

The scans were conducted with a 3T Siemens MAGNETOM Trio scanner with a 32-channel head array coil at the USC Dana and David Dornsife Neuroimaging Center. T1-weighted 3D structural MRI brain scans were acquired pre and post-intervention using a magnetization prepared rapid acquisition gradient echo (MPRAGE) sequence with the parameters of TR = 2300 ms, TE = 2.26 ms, slice thickness = 1.0 mm, flip angle = 9°, field of view = 256 mm, and voxel size = 1.0 x 1.0 mm, with 175 volumes collected (4:44 min).

### 2.4. Data Processing and Analysis

#### 2.4.1. Texture Analysis

Among the 106 younger adults and 56 older adults who completed all 7 weeks of the study, 100 younger adults and 51 older adults finished pre-intervention and post-intervention MRI sessions.

We selected 10 regions of interest (ROIs) based on prior evidence that texture-based measures derived from T1-weighted MRI are sensitive to Alzheimer’s disease–related neurodegenerative processes and radiographic pathology, and may capture microstructural changes beyond what is reflected by regional volume alone (Kwon et al., 2023; S. Lee et al., 2020, 2021). The ROIs included the four Alzheimer’s disease signature cortical regions (entorhinal, inferior temporal, middle temporal, and fusiform cortex) and the hippocampus, each examined separately for the left and right hemispheres (Figure 1). ROI masks were generated using FreeSurfer 7.3.1 (surfer.nmr.mgh.harvard.edu/) following previously described pipelines (Kwon et al., 2023; S. Lee et al., 2021). ROI images were created by applying these masks to each participant’s native T1-weighted scans while retaining original intensity values.

**Fig 1.**
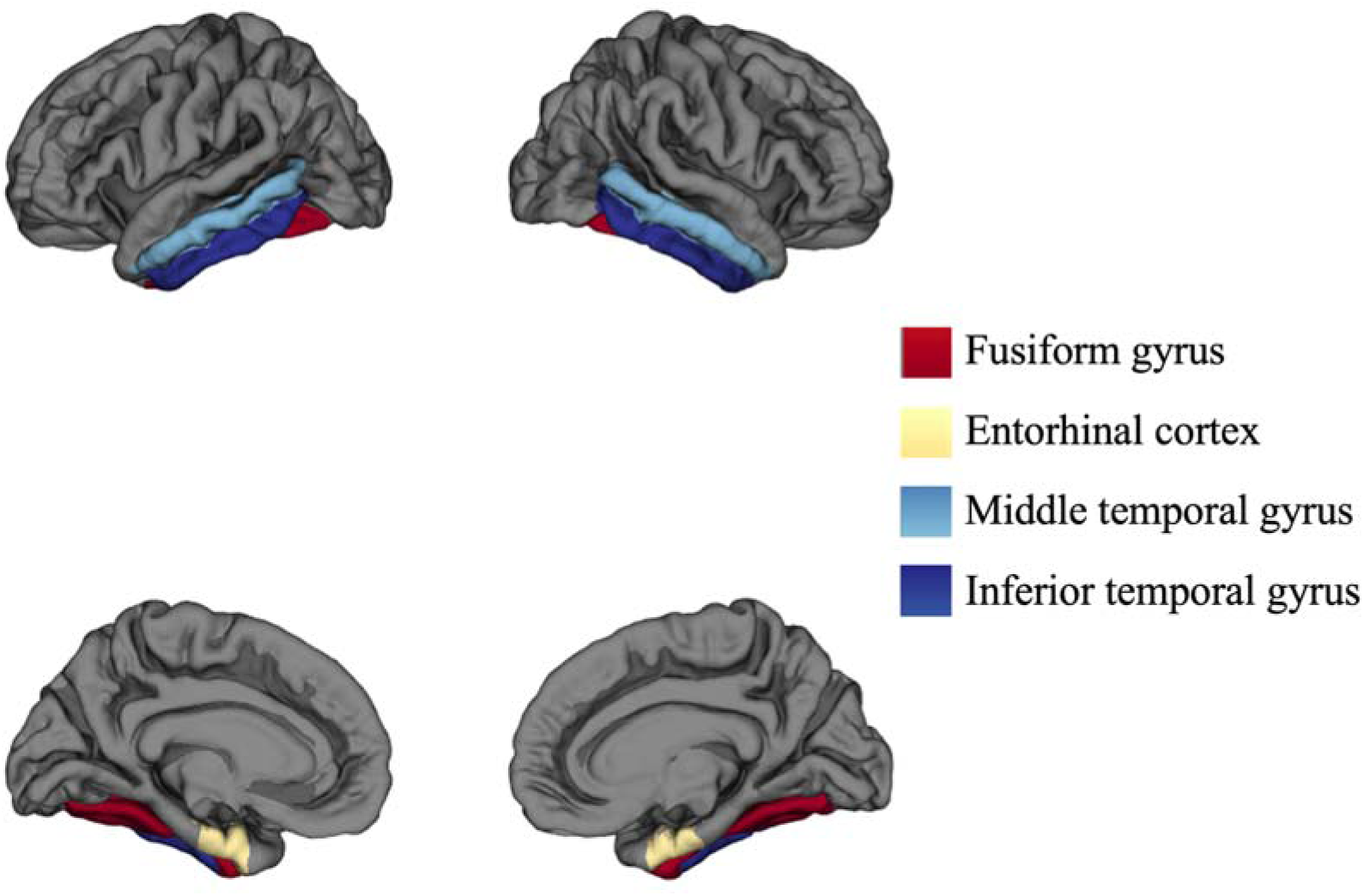
Map of the four Alzheimer signature regions based on FreeSurfer cortical labels, shown on the pial surface of the left (left) and right (right) hemispheres for lateral (top) and medial (bottom) views.

Prior to feature extraction, ROI images underwent several preprocessing steps consistent with established hippocampal texture analysis pipelines. First, histogram normalization was applied by excluding voxels with intensity values outside μ ± 3σ (where μ and σ represent the mean and standard deviation of voxel intensities within each ROI), to mitigate partial volume effects. Next, voxel intensities were normalized relative to the mean cerebrospinal fluid (CSF) intensity extracted from the lateral ventricles, reducing interindividual variability in intensity scaling. Finally, voxel intensities were quantized into 32 gray levels to stabilize the estimation of co-occurrence matrices. No spatial normalization was applied in order to preserve native intensity distributions.

Texture features were computed using three-dimensional gray-level co-occurrence matrix (3D-GLCM) analysis, following previously published methods shown to be sensitive to Alzheimer’s disease–related radiographic pathology, including amyloid- and tau-associated changes (Lee et al., 2021). GLCMs were generated for voxel pairs at a distance of one voxel across 13 orientations in three-dimensional space, incorporating both within-slice and cross-slice relationships. Each GLCM was normalized and subsequently averaged across orientations to yield a representative matrix for each ROI.

From these matrices, we extracted 22 Haralick texture features, including entropy, contrast, autocorrelation, cluster shade, and additional indices capturing complementary aspects of spatial intensity variation. Feature extraction was implemented using analysis scripts adapted from previously published work (S. Lee et al., 2020, 2021) and applied consistently across all ROIs and participants. All 22 features were computed for each of the 10 ROIs, and the full list of features and corresponding results are provided in Supplementary Table S1.

#### 2.4.2. Statistical analysis on texture features

We performed outlier detection on the texture indices using a multivariate approach in the combined pre- and post-training data space. All 22 texture features from the ADSC region at both time points (pre- and post-training) were concatenated into a single 44-dimensional feature vector for each participant and standardized using z-score normalization. We then applied principal component analysis (PCA) to this combined feature space and conducted outlier detection using Mahalanobis distance. Specifically, Mahalanobis distances were calculated for each participant based on the first two principal components of the combined PCA space. The Mahalanobis distance quantifies how far each observation is from the multivariate centroid, accounting for correlations among variables. Participants with Mahalanobis distances exceeding the 99th percentile were flagged as multivariate outliers and excluded from subsequent analyses. This procedure resulted in the exclusion of 2 participants (original N = 151, final N = 149).

Following outlier removal, all 22 Haralick texture features were standardized by converting each measure to z-scores. Explanatory factor analysis was then conducted separately for each ROI to reduce dimensionality and derive composite factor scores. IBM SPSS Statistics version 28 was used for all factor analyses.

For each ROI, factor analysis was first performed on pre-intervention data. Bartlett’s Test of Sphericity and the Kaiser–Meyer–Olkin (KMO) measure of sampling adequacy confirmed the suitability of the data for factor analysis. Principal component analysis (PCA) with varimax rotation was applied, and factors with Eigenvalues greater than 1 were retained. The resulting factor structure from the pre-intervention data was then used to compute factor scores for the post-intervention data by applying the same loading matrix, thereby ensuring comparability of factor scores across time points.

From these analyses, a reduced set of factor scores was obtained for each ROI. To test the effects of time (pre, post), training condition (Osc+, Osc-), and age group (younger adults, older adults) on the derived factor scores, we conducted a 2 × 2 × 2 mixed-design analysis using linear mixed-effects models (LMMs) with random intercepts for participants. Rather than employing additional outlier exclusion procedures based on the factor score distributions, we adopted a robust statistical approach to accommodate potential influential observations. Specifically, we used cluster-robust variance estimation with bias-reduced linearization (CR2 method; Bell & McCaffrey, 2002; Pustejovsky & Tipton, 2018) implemented in the clubSandwich R package (Pustejovsky, 2020). This method provides valid inference in the presence of heteroscedasticity and within-cluster dependence inherent in repeated-measures designs, while maintaining robustness against potential outliers. All hypothesis tests (F tests for omnibus effects and t tests for specific contrasts) were conducted using these cluster-robust standard errors, with participants serving as the clustering unit. Statistical significance was determined at α = .05.s determined at α = .05.

#### 2.4.3. Texture–Plasma biomarker PLSC

We conducted partial least squares correlation (PLSC) analyses to examine multivariate associations between training-related changes in plasma Alzheimer’s disease–related biomarkers and regional texture features derived from left and right ADSC and hippocampal regions. This multivariate approach is well suited for neuroimaging–biomarker analyses because it accommodates multicollinearity among predictors and identifies latent dimensions linking plasma biomarker profiles with distributed patterns of structural texture variation (Krishnan et al., 2011).

The X matrix comprised post–pre difference scores of plasma biomarkers, including Aβ40, Aβ42, total tau, phosphorylated tau (p-tau), and the Aβ42/40 ratio. On the Y side, the matrix contained post–pre difference scores of 22 Haralick texture features derived from the left and right ADSC and hippocampus. All variables were standardized (z-scored) prior to entry into the analysis.

PLSC identifies latent variables (LVs) that capture maximal shared covariance between two sets of measures. Statistical significance of each LV was assessed using permutation testing (n = 1000), in which the rows of the Y matrix were randomly permuted to generate a null distribution of singular values. The p-value for each LV was defined as the proportion of permuted singular values exceeding the observed singular value. Reliability of individual biomarker and texture contributions was evaluated using bootstrap resampling (n = 1000) with replacement. Bootstrap ratios (salience divided by its standard error) were computed, with |BSR| > 1.96 indicating statistically reliable contributors.

This multivariate framework enabled identification of coordinated patterns linking regional microstructural texture reorganization with concurrent changes in plasma amyloid-β and tau-related biomarkers that may not be evident in univariate analyses.

## 3. Results

### 3.1 Sample Characteristics

Baseline demographic characteristics of the participants by age group and condition are summarized in Table 1.

**Table 1.** Basic participant characteristics for each condition in each age group. Means and standard deviation (in parenthesis) are provided. (N=151).

|  | Younger Adults |  | Older Adults |  |
| --- | --- | --- | --- | --- |
|  | Osc+ (N=52) | Osc- (N=48) | Osc+ (N=26) | Osc- (N=25) |
| Age (years) | 22.83 (2.40) | 22.73 (3.17) | 65.38 (7.88) | 64.60 (5.48) |
|  | All: 22.78 (2.80) |  | All: 65.00 (6.75) |  |
| Sex | 26 F / 26 M | 24 F / 24 M | 19 F / 7 M | 18 F / 7 M |
|  | All: 50 F / 50 M |  | All: 37 F / 14 M |  |
| Education | 16.13 (1.76) | 15.83 (2.56) | 17.12 (2.50) | 16.26 (2.39) |
|  | All: 15.98 (2.18) |  | All: 16.70 (2.45) |  |

### 3.2 Intervention Effects on Composite Texture Indices

In the ADSC region, three factor scores were derived from principal component analysis of texture indices. We conducted a 2 (time: pre, post) × 2 (condition: Osc+, Osc-) × 2 (age group: younger adults, older adults) mixed-design analysis using linear mixed-effects models with cluster-robust variance estimation (N = 149, df = 141 for F-tests). For Factors 1 and 2, no significant two-way or three-way interactions emerged (all *Fs* < 2.65, *ps* > .10, η*²p* < .018).

Factor 3’s Time × Condition interaction also did not reach the conventional levels of significance, *F*(1, 141) = 3.91, *p* = .050, η*²p* = .027 (see Figure 2a). Although neither the Osc-group (*b* = −0.29, *SE* = 0.21, *t*(145.0) = −1.39, *p* = .166, *d* = −0.12) nor the Osc+ group (*b* = 0.27, *SE* = 0.19, *t*(145.0) = 1.41, *p* = .162, *d* = 0.12) exhibited significant simple effects, the significant interaction emerged from opposing patterns of change between conditions. The Time × Condition × Age Group three-way interaction was not significant, *F*(1, 141) = 2.96, *p* = .088, η*²p* = .021. Exploratory analyses examining the two-way interaction separately for each age group revealed that the pattern was primarily driven by older adults, *F*(1, 141) = 3.58, *p* = .061, η*²p* = .025, whereas younger adults showed no such interaction, *F*(1, 141) = 0.37, *p* = .545, η*²p* = .003 (Figure 2b). Across all three factors, significant main effects of age group emerged for Factors 1 and 2 (*F*(1, 141) = 56.97, *p* < .001, η*²p* = .288; *F*(1, 141) = 15.78, *p* < .001, η*²p* = .101), with older adults showing lower scores than younger adults. No other main effects or interactions reached significance (all *ps* > .10).

**Fig 2.**
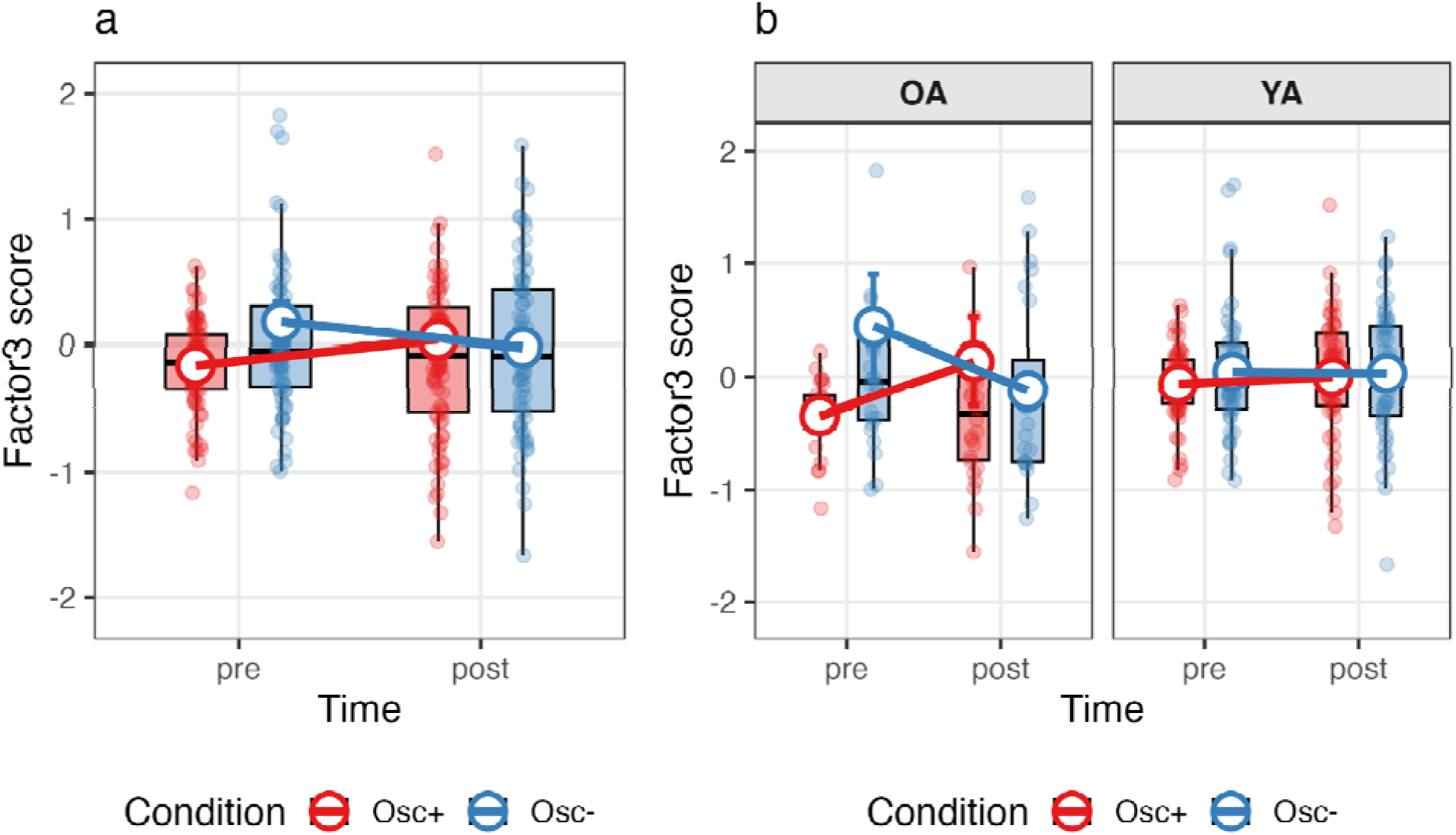
Time × Condition Interaction for Texture Factor 3 in the ADSC Region. (a) Overall Time × Condition interaction. (b) Time × Condition interaction by age group (YA = younger adults; OA = older adults). Boxplots (quartiles and 1.5 × IQR whiskers) and group means ± SE (large circles connected by red and blue bold lines) are displayed. All statistics computed from N = 149 participants after exclusion of multivariate outliers (Mahalanobis distance). The y-axis range was constrained to boxplot whisker limits to minimize visual distortion from extreme values. Three data points fall outside the displayed range (1 at pre-test, 2 at post-test) but were fully included in all statistical analyses. Osc+ = oscillation-increase group; Osc-= oscillation-decrease group.

In the hippocampus, four factor scores were derived from principal component analysis, accounting for 97% of the total variance. Using the same mixed-design analytical approach (N = 149, df = 141), Factors 1, 2, and 3 showed no significant two-way or three-way interactions (all *Fs* < 3.18, ps > .076, η*²p* < .023). Factor 4 revealed a significant Time × Condition × Age Group three-way interaction, *F*(1, 141) = 4.21, *p* = .042, η*²p* = .029 (see Figure 3). Decomposition of this interaction by age group revealed a significant Time × Condition interaction in older adults, *F*(1, 141) = 10.39, *p* < .001, η*²p* = .069, but not in younger adults, *F*(1, 141) = 0.72, *p* = .397, η*²p* = .005. Within older adults, the Osc-group showed a decrease in Factor 4 scores (*b* = −0.56, *SE* = 0.40, *t*(145.0) = −1.40, *p* = .165, *d* = −0.12) while the Osc+ group exhibited an increase (*b* = 0.48, *SE* = 0.37, *t*(145.0) = 1.28, *p* = .204, *d* = 0.11), though neither simple effect was independently significant. The overall Time × Condition interaction (collapsed across age groups) was not significant, *F*(1, 141) = 3.18, *p* = .077, η*²p* = .022. No significant main effects emerged for any factor (all *Fs* < 2.40, *ps* > .10).

**Fig 3.**
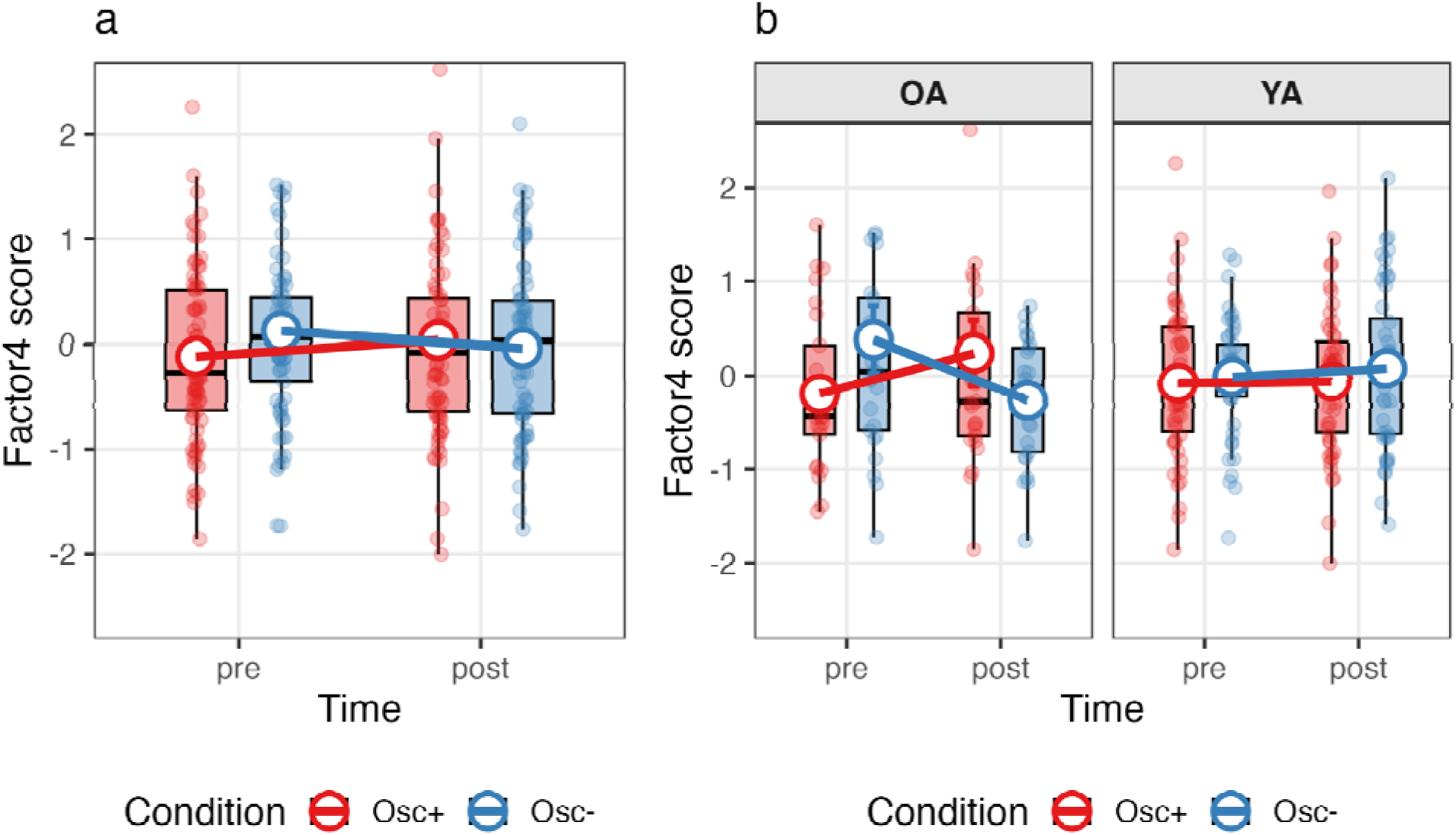
Time × Condition × Age Group Interaction for Texture Factor 4 in the Hippocampus. (a) Overall Time × Condition interaction. (b) Time × Condition interaction by age group (YA = younger adults; OA = older adults). Boxplots (quartiles and 1.5 × IQR whiskers) and group means ± SE (large circles connected by red and blue bold lines) are displayed. All statistics computed from N = 149 participants after exclusion of multivariate outliers (Mahalanobis distance). The y-axis range was constrained to boxplot whisker limits to minimize visual distortion from extreme values. Data points falling outside the displayed range (1 at pre-test, 2 at post-test) were fully included in all statistical analyses. Osc+ = oscillation-increase group; Osc-= oscillation-decrease group.

### 3.3 Intervention Effects on Individual Texture Metrics

To provide a more precise interpretation of the factor-level effects, we conducted follow-up analyses on individual texture indices that showed the strongest factor loadings on the significant factors. For ADSC Factor 3, we examined T1 (Uniformity; loading = .986) and T2 (Entropy; loading = -.940); for hippocampus Factor 4, we examined T1 (Uniformity; loading = .951) and T2 (Entropy; loading = -.894). These indices represent opposite aspects of texture regularity: higher uniformity and lower entropy both indicate better organized patterns.

For the ADSC region, both indices showed significant Time × Condition interactions, T1: *F*(1, 141) = 4.48, *p* = .036, η*²p* = .031, Figure 4a; T2: *F*(1, 141) = 4.91, *p* = .028, η*²p* = .034, (Figure 4c). Although the three-way interactions were not significant (T1: *F*(1, 141) = 3.02, *p* = .084, η*²p* = .021; T2: *F*(1, 141) = 4.42, *p* = .037, η*²p* = .030), exploratory age-stratified analyses revealed that the effects were specific to older adults (Figure 4b, 4d). For T1, older adults showed a Time × Condition interaction, *F*(1, 141) = 3.91, *p* = .050, η*²p* = .027, which did not reach conventional levels of statistical significance, and no interaction was observed in younger adults, *F*(1, 141) = 0.71, *p* = .400, η*²p* = .005. For T2, the interaction was significant in older adults (*F*(1, 141) = 4.92, *p* = .028, η*²p* = .034) but not younger adults (*F*(1, 141) = 0.06, *p* = .804, η*²p* < .001). The convergent pattern of increased uniformity and decreased entropy in older adults receiving Osc+ training indicated improved texture regularity, confirming the Factor 3 effects.

**Fig 4.**
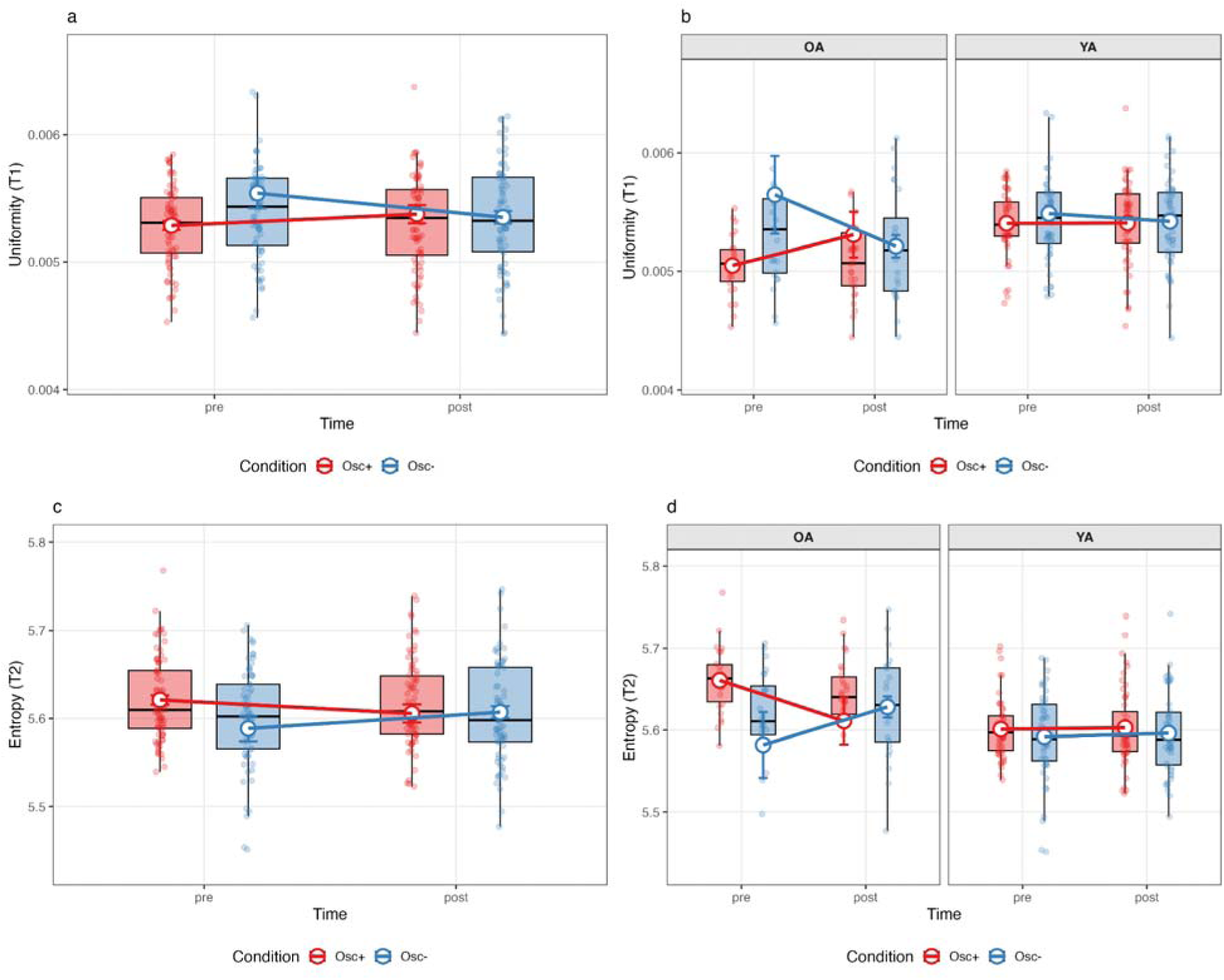
Time × Condition Interactions for Individual Texture Indices in the ADSC (Uniformity: panels A-B; Entropy: panels C-D). Panel A shows the two-way Time×Condition interaction for uniformity, with the decrease condition exhibiting increased uniformity from pre-to post-intervention. Panel B displays the three-way interaction (Time×Condition×Age Group) for uniformity, showing that the pattern was primarily driven by older adults. Panel C shows the two-way Time×Condition interaction for entropy, with the decrease condition showing reduced entropy. Panel D displays the three-way interaction for entropy, revealing significant effects in older adults but not younger adults. The complementary patterns for uniformity (increase) and entropy (decrease) both indicate increased textural regularity following HRV-decrease training in older adults’ ADSC. Error bars represent standard errors. Osc+ = HRV-increase training; Osc-= HRV-decrease training; OA = older adults; YA = younger adults. Data points beyond 1.5 IQR from the box plot whiskers were excluded from visualization (T1: n=6; T2: n=3).

For the hippocampus, both indices showed significant three-way interactions (T1: *F*(1,141) = 4.22, *p* = .042, η*²p* = .029, Figure 5b; T2: *F*(1,141) = 4.06, *p* = .046, η*²p* = .028, Figure 5d). Follow-up analyses revealed that the simple two-way Time × Condition interactions within older adults approached significance for both indices (*F*(1,141) = 3.57, p = .061, η*²p* = .025 for each; Figure 5a, 5c). The opposite directional patterns between conditions for uniformity and entropy, specific to older adults, suggested differential effects of HRV training on hippocampal texture, contrasting with the texture-enhancing effects observed in ADSC following HRV-increase training.

**Fig 5.**
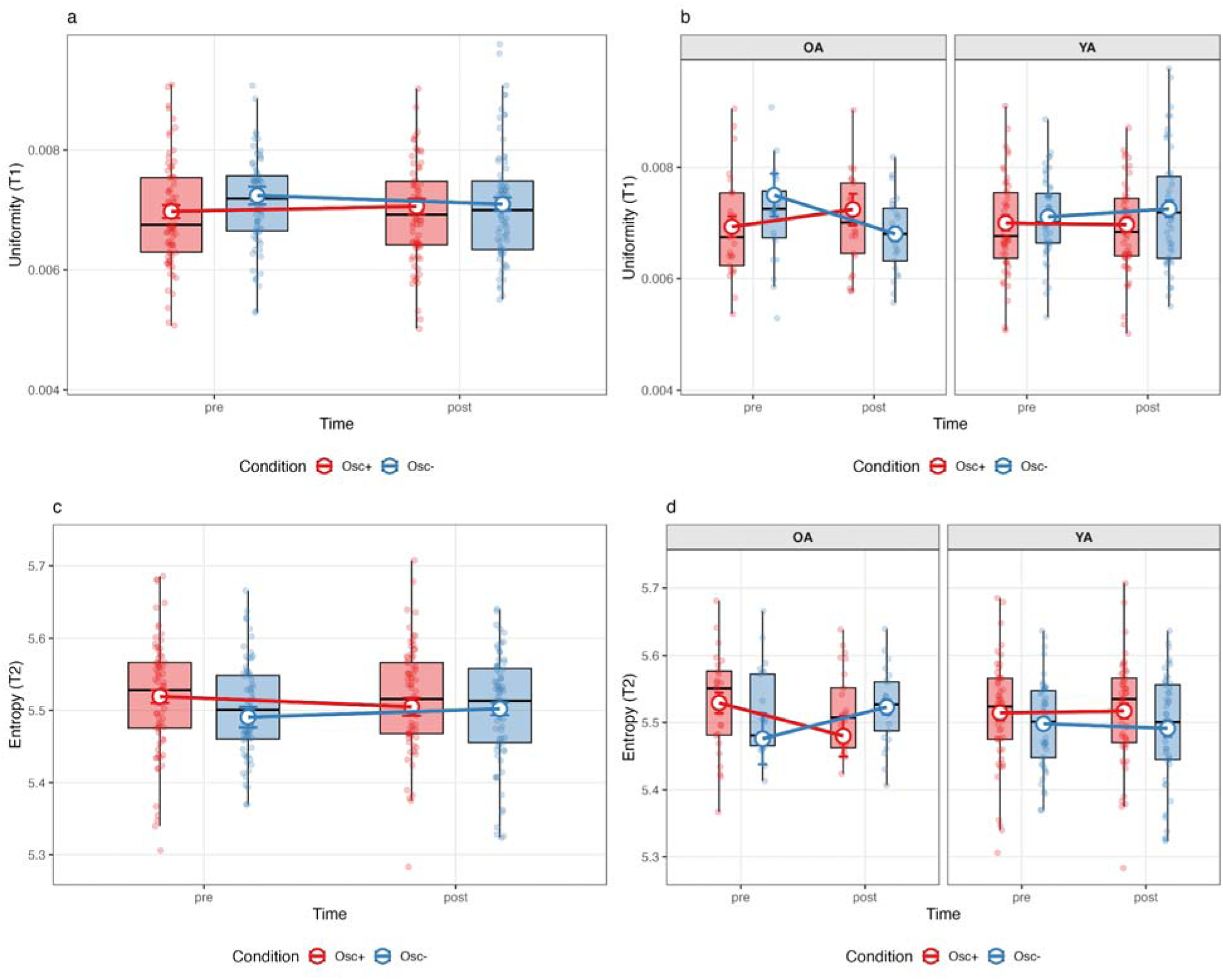
Time × Condition × Age Group Interactions for Individual Texture Indices in the Hippocampus (Uniformity: panels A-B; Entropy: panels C-D). Panel A shows the two-way Time×Condition interaction for uniformity. Panel B displays the three-way interaction, revealing that the decrease condition led to reduced uniformity specifically in older adults. Panel C shows the two-way Time×Condition interaction for entropy. Panel D displays the three-way interaction for entropy, showing increased entropy following decrease training in older adults. The complementary patterns for uniformity (decrease) and entropy (increase) both indicate increased textural heterogeneity in older adults’ hippocampus following HRV-decrease training. Error bars represent standard errors. Osc+ = HRV-increase training; Osc-= HRV-decrease training; OA = older adults; YA = younger adults. Data points beyond 1.5 IQR from the box plot whiskers were excluded from visualization (T1: n=5; T2: n=3).

### 3.4 Hemispheric and Subregional Time × Condition Effects in the ADSC and Hippocampus

To further characterize the hemispheric and subregional distribution of the whole-region effects observed in the ADSC and hippocampus, we conducted follow-up analyses on individual texture indices that showed the strongest loadings on the composite factors. Across all factors identified in the preceding composite analyses, T1 (Uniformity) and T2 (Entropy) exhibited the highest absolute factor loadings, indicating that these indices primarily contributed to the composite effects. We therefore examined hemispheric and subregional Time × Condition interactions for T1 and T2 to determine whether the whole-region effects were localized to specific subregions or hemispheres. Full results for all subregions, age-group–specific effects, and three-way interactions are summarized in Table 2.

**Table 2.** Time × Condition Interaction Effects for Individual Texture Indices (T1: Uniformity; T2: Entropy) Across Subregions and Hemispheres.

| Hemi-sphere | Region | Texture | Time × Condition | Time × Condition (OA) | Time × Condition (YA) | Time × Cond × Age group |
| --- | --- | --- | --- | --- | --- | --- |
| Left | entorhinal | T1 | $F = 2.49$<br>($p = .117$) | $F = 2.30$<br>($p = .132$) | $F = 0.19$<br>($p = .661$) | $F = 1.78$<br>($p = .184$) |
| | | T2 | $F = 2.68$<br>($p = .104$) | $F = 2.90$<br>($p = .091$) | $F = 0.01$<br>($p = .938$) | $F = 2.53$<br>($p = .114$) |
| | fusiform | T1 | $F = 2.15$<br>($p = .145$) | $F = 1.63$<br>($p = .204$) | $F = 1.44$<br>($p = .232$) | $F = 1.10$<br>($p = .295$) |
| | | T2 | $F = 1.48$<br>($p = .226$) | $F = 1.02$<br>($p = .314$) | $F = 1.19$<br>($p = .278$) | $F = 0.58$<br>($p = .448$) |
| | inferior temporal | T1 | $F = 5.86^*$<br>($p = .017$) | $F = 3.61$<br>($p = .059$) | $F = 5.15^*$<br>($p = .025$) | $F = 1.57$<br>($p = .212$) |
| | | T2 | $F = 4.82^*$<br>($p = .030$) | $F = 3.41$<br>($p = .067$) | $F = 2.86$<br>($p = .093$) | $F = 1.93$<br>($p = .167$) |
| | mid temporal | T1 | $F = 7.77^{**}$<br>($p = .006$) | $F = 5.24^*$<br>($p = .024$) | $F = 5.41^*$<br>($p = .021$) | $F = 2.74$<br>($p = .100$) |
| | | T2 | $F = 6.40^*$<br>($p = .013$) | $F = 4.66^*$<br>($p = .033$) | $F = 3.34$<br>($p = .070$) | $F = 2.76$<br>($p = .099$) |
| | hippocampus | T1 | $F = 1.86$<br>($p = .175$) | $F = 2.94$<br>($p = .089$) | $F = 0.73$<br>($p = .393$) | $F = 3.59$<br>($p = .060$) |
| | | T2 | $F = 1.65$<br>($p = .201$) | $F = 2.93$<br>($p = .089$) | $F = 0.88$<br>($p = .351$) | $F = 3.72$<br>($p = .056$) |
| Right | entorhinal | T1 | $F = 4.97^*$<br>($p = .027$) | $F = 3.72$<br>($p = .056$) | $F = 1.80$<br>($p = .182$) | $F = 2.16$<br>($p = .143$) |
| | | T2 | $F = 4.92^*$<br>($p = .028$) | $F = 3.58$<br>($p = .060$) | $F = 1.82$<br>($p = .180$) | $F = 1.90$<br>($p = .170$) |
| | fusiform | T1 | $F = 3.90$<br>($p = .050$) | $F = 3.98^*$<br>($p = .048$) | $F = 0.01$<br>($p = .938$) | $F = 3.79$<br>($p = .053$) |
| | | T2 | $F = 4.31^*$<br>($p = .040$) | $F = 4.41^*$<br>($p = .038$) | $F = 0.03$<br>($p = .867$) | $F = 3.97^*$<br>($p = .048$) |
| | inferior temporal | T1 | $F = 3.15$<br>($p = .078$) | $F = 3.27$<br>($p = .073$) | $F = 0.01$<br>($p = .929$) | $F = 2.99$<br>($p = .086$) |
| | | T2 | $F = 2.70$<br>($p = .102$) | $F = 3.15$<br>($p = .078$) | $F = 0.07$<br>($p = .792$) | $F = 3.18$<br>($p = .077$) |
| | mid temporal | T1 | $F = 4.95^*$<br>($p = .028$) | $F = 2.66$<br>($p = .105$) | $F = 3.67$<br>($p = .058$) | $F = 0.65$<br>($p = .423$) |
| | | T2 | $F = 2.48$<br>($p = .117$) | $F = 1.64$<br>($p = .203$) | $F = 1.10$<br>($p = .295$) | $F = 0.65$<br>($p = .422$) |
| | hippocampus | T1 | $F = 4.03^*$<br>($p = .047$) | $F = 2.37$<br>($p = .126$) | $F = 3.91^*$<br>($p = .050$) | $F = 0.92$<br>($p = .339$) |
| | | T2 | $F = 3.99^*$<br>($p = .048$) | $F = 2.30$<br>($p = .132$) | $F = 3.29$<br>($p = .072$) | $F = 0.79$<br>($p = .376$) |

*Note.* $F$ statistics and $p$ values are shown for each interaction term. *Time × Condition* (OA) and *Time × Condition* (YA) indicate age-group-specific simple effects within older adults (OA) and younger adults (YA), respectively. T1 corresponds to Uniformity and T2 to Entropy. ADSC = Alzheimer's disease-signature cortical region; hippo = hippocampus. Significant effects are indicated by \* ( $p < .05$ ) and \*\* ( $p < .01$ ). \* did not remain significant after multiple comparison corrections using the Benjamini-Hochberg procedure.

At the uncorrected level, several subregional Time x Condition effects were observed, particularly in the lateral temporal cortical regions and the right hippocampus. However, none of these subregional effects remained significant after Benjamini-Hochberg correction for multiple comparisons.

Within the ADSC, the strongest nominal effects were observed in the left inferior and middle temporal cortices. In the left inferior temporal region, Time × Condition interactions were detected for both T1 and T2, with some age-group–specific effects in younger adults for T1 and weaker effects for T2. In the left middle temporal region, nominal Time × Condition interactions were observed for both T1 and T2, with age-group-specific follow-up effects suggesting that the pattern may have been more pronounced in younger adults for T1 and in older adults for T2 In the right ADSC, nominal Time × Condition interactions emerged in the entorhinal cortex for both T1 and T2 and in the middle temporal region for T1. A three-way Time × Condition × Age group interaction was observed in the right fusiform gyrus for T2, with Time x Condition effects suggesting that the pattern was driven primarily by older adults.

In the hippocampus, hemispheric differences were also evident at the uncorrected level. Nominal Time × Condition interactions were detected in the right hippocampus for both T1 and T2, with an additional age group–specific effect for T1 in younger adults, whereas only trend-level three-way interactions were observed in the left hippocampus. Together, these findings suggest that the whole-region effects in the ADSC may be distributed across lateral temporal subregions, whereas hippocampal effects may be more lateralized to the right hemisphere, with limited age-group–dependent modulation.

### 3.5 Brain–plasma AD biomarker associations across ADSC and hippocampal texture changes

To examine whether training-related texture changes across ADSC and hippocampal regions were associated with concurrent changes in plasma Alzheimer’s disease–related biomarkers, we conducted partial least squares correlation (PLSC) analyses using post–pre difference scores for multiple bilateral regions of interest. Several regions showed significant brain–plasma coupling. We first describe results for the right entorhinal cortex (Figure 6).

**Fig 6.**
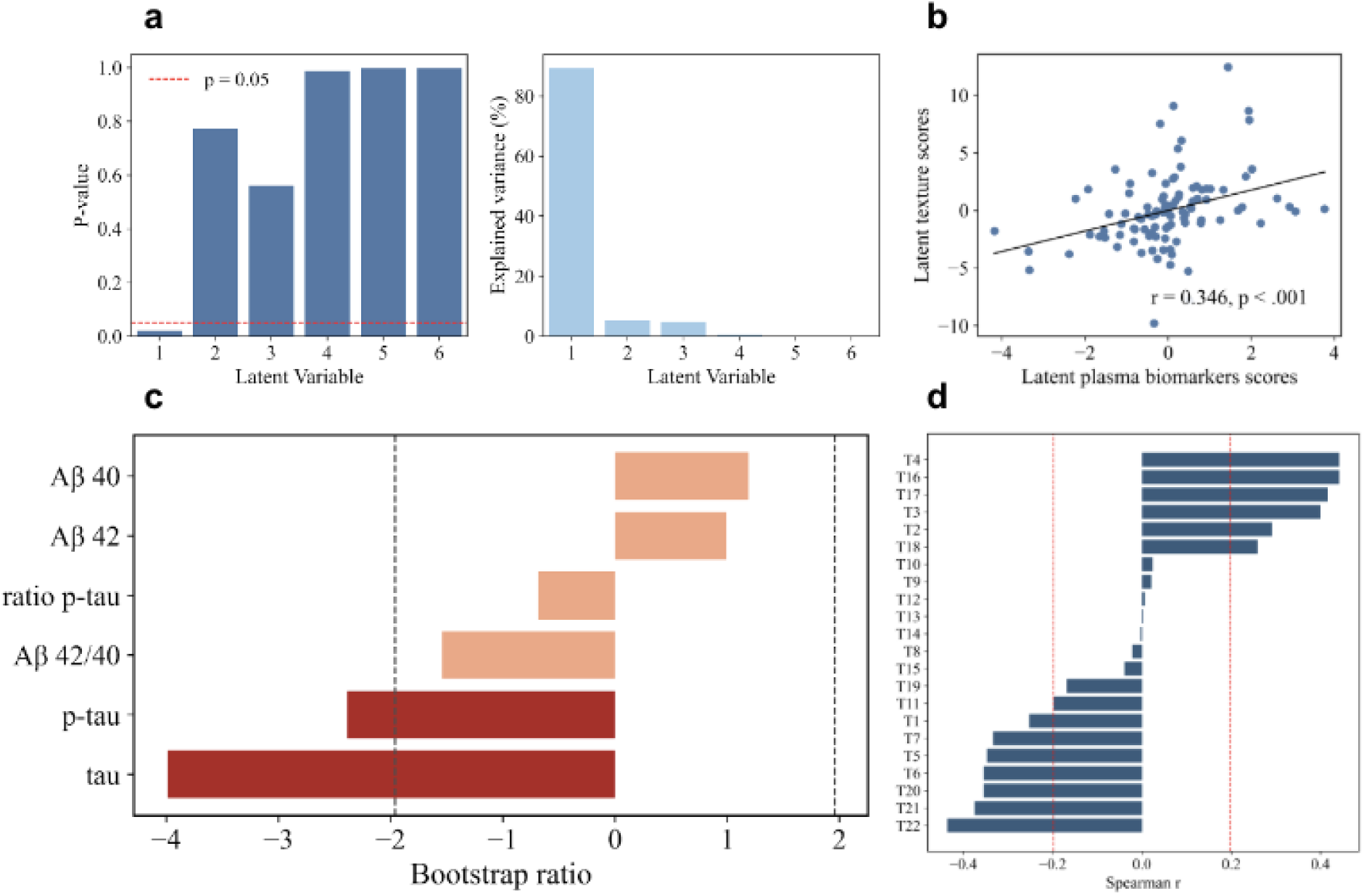
Partial least squares correlation (PLSC) results for latent variable 1 (LV1): Associations between training-related changes in right entorhinal textures and changes in plasma AD-related biomarkers. (a) P-values of latent variables, showing that only LV1 reached statistical significance (p < .05). (b) Correlation between latent texture scores and latent plasma biomarker scores. (c) Bootstrap ratios for plasma AD-related biomarker variables (X-side), reflecting the contributions of biomarker changes to LV1. (d) Spearman correlations between texture measures and plasma biomarker latent scores (Y-side). T1 = Uniformity/Energy/Angular Second Moment, T2 = Entropy, T3 = Dissimilarity, T4 = Contrast/Inertia, T5 = Inverse Difference, T6 = Correlation, T7 = Homogeneity/Inverse Difference Moment, T8 = Autocorrelation, T9 = Cluster Shade, T10 = Cluster Prominence, T11 = Maximum Probability, T12 = Sum of Squares, T13 = Sum Average, T14 = Sum Variance, T15 = Sum Entropy, T16 = Difference Variance, T17 = Difference Entropy, T18 = Information Measure of Correlation 1 (IMC1), T19 = Information Measure of Correlation 2 (IMC2), T20 = Maximal Correlation Coefficient, T21 = Inverse Difference Normalized (IDN), T22 = Inverse Difference Moment Normalized (IDMN).

The analysis revealed one significant latent variable (LV1; *p* < .05), accounting for 89.41% of the explained covariance, indicating a robust multivariate association between training-related changes in entorhinal texture features and concurrent changes in plasma AD-related biomarkers (Figure 6a). Latent brain and plasma biomarker scores were moderately correlated (r = .346, *p* < .001; Figure 6b), reflecting coordinated brain–plasma changes following training. On the plasma side, bootstrap resampling showed that tau average difference (BSR = −3.991) and p-tau average difference (BSR = −2.386) were the most reliable contributors to LV1 (Figure 6c), indicating that tau-related biomarkers primarily drove the plasma profile.

On the texture side, Spearman correlations between individual texture features and the latent brain score revealed a coherent pattern of entorhinal microstructural change (Figure 6d). Features reflecting greater tissue homogeneity and spatial regularity—including inverse difference–based measures, homogeneity, correlation, and inverse difference moment normalized—were positively associated with the latent brain score, whereas features capturing increased heterogeneity or disorder—including entropy-, variance-, and contrast-related measures—were negatively associated. Interpretation of directionality accounted for the feature-specific biological meaning of T1-weighted texture measures (Supplementary Table S1).

Together, these findings indicate that training-related increases in texture features indexing greater tissue regularity, accompanied by concurrent reductions in disorder-related features, were associated with decreases in plasma tau-related biomarkers. This multivariate profile suggests that microstructural reorganization of the entorhinal cortex following training is linked to biologically favorable changes in tau pathology.

We next describe results for the right fusiform region (Figure 7). The analysis again identified a single significant latent variable (LV1; *p* < .05), accounting for 93.05% of the explained covariance, demonstrating a strong multivariate coupling between fusiform texture changes and plasma AD-related biomarkers (Figure 7a). Latent brain and plasma biomarker scores were significantly correlated (*r* = .329, *p* < .001; Figure 7b), indicating coordinated brain–plasma modulation following training. Bootstrap analyses showed that p-tau ratio difference (BSR = −2.682) and p-tau average difference (BSR = −3.189) were the dominant contributors on the plasma side (Figure 7c).

**Fig 7.**
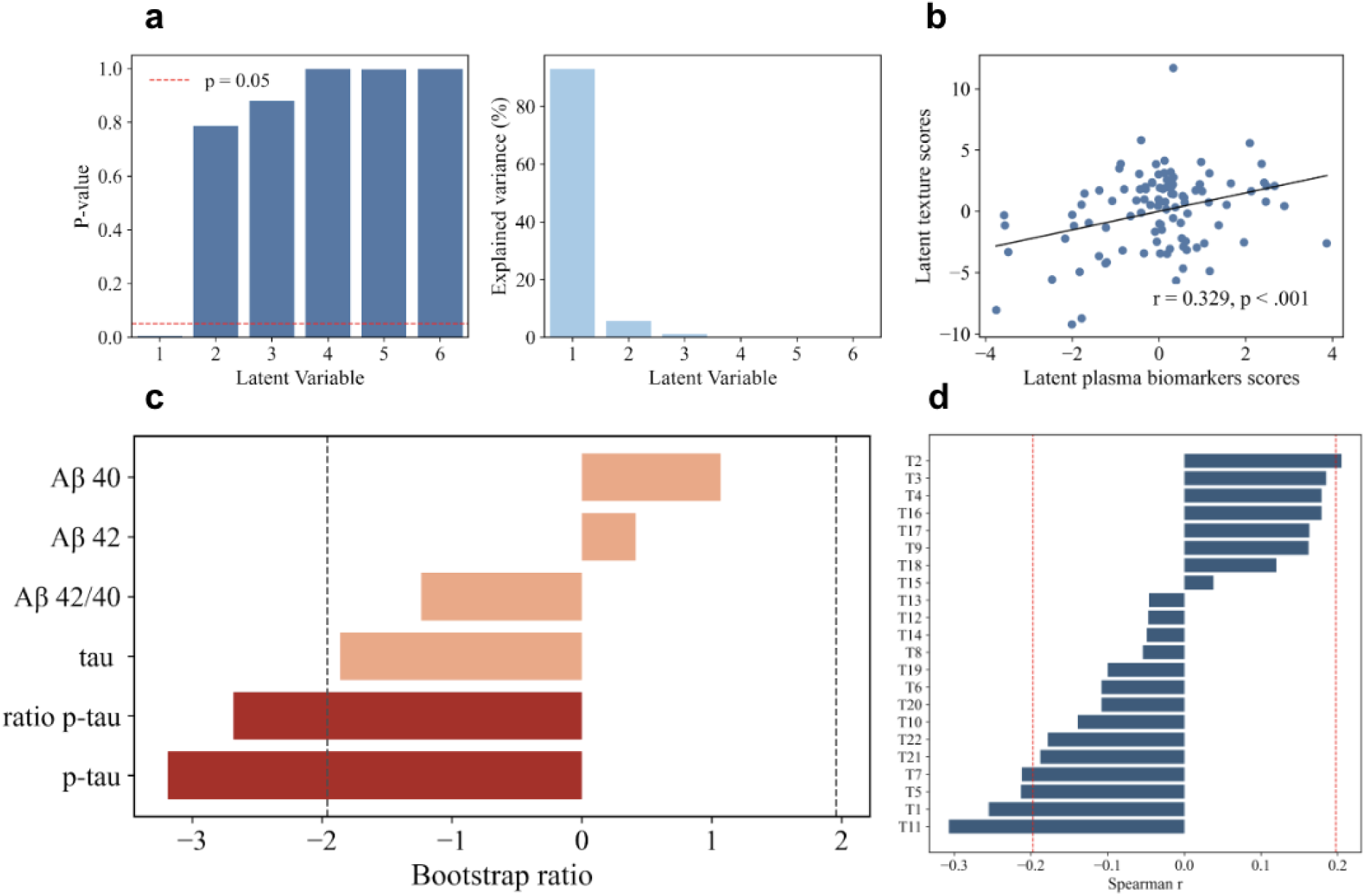
Partial least squares correlation (PLSC) results for latent variable 1 (LV1): Associations between training-related changes in right fusiform textures and changes in plasma AD-related biomarkers. (a) P-values of latent variables, showing that only LV1 reached statistical significance (p < .05). (b) Correlation between latent texture scores and latent plasma biomarker scores. (c) Bootstrap ratios for plasma AD-related biomarker variables (X-side), reflecting the contributions of biomarker changes to LV1. (d) Spearman correlations between texture measures and plasma biomarker latent scores (Y-side). T1 = Uniformity/Energy/Angular Second Moment, T2 = Entropy, T3 = Dissimilarity, T4 = Contrast/Inertia, T5 = Inverse Difference, T6 = Correlation, T7 = Homogeneity/Inverse Difference Moment, T8 = Autocorrelation, T9 = Cluster Shade, T10 = Cluster Prominence, T11 = Maximum Probability, T12 = Sum of Squares, T13 = Sum Average, T14 = Sum Variance, T15 = Sum Entropy, T16 = Difference Variance, T17 = Difference Entropy, T18 = Information Measure of Correlation 1 (IMC1), T19 = Information Measure of Correlation 2 (IMC2), T20 = Maximal Correlation Coefficient, T21 = Inverse Difference Normalized (IDN), T22 = Inverse Difference Moment Normalized (IDMN).

Texture–latent score associations revealed a structured fusiform pattern characterized by enhanced regularity and reduced heterogeneity (Figure 7d). In particular, inverse difference–based measures, homogeneity, and maximal probability were positively associated with the latent brain score, whereas entropy- and variance-related features showed negative associations, consistent with improved microstructural organization. Directional interpretation was guided by feature-specific biological meaning (Supplementary Table S1).

These results indicate that training-related fusiform microstructural reorganization, marked by increasing spatial regularity and decreasing disorder, is coupled with reductions in plasma p-tau–related biomarkers, suggesting coordinated cortical–molecular plasticity.

Finally, we report results for the left hippocampus (Figure 8). A single significant latent variable (LV1; *p* < .05) explained 51% of the covariance between hippocampal texture changes and plasma biomarker changes (Figure 8a). Latent brain and plasma biomarker scores were moderately correlated (*r* = .349, *p* < .001; Figure 8b), representing the strongest brain–plasma coupling observed across the examined regions. On the plasma side, bootstrap resampling identified p-tau ratio difference (BSR = −2.087) and p-tau average difference as the most reliable contributors to LV1 (Figure 8c).

**Fig 8.**
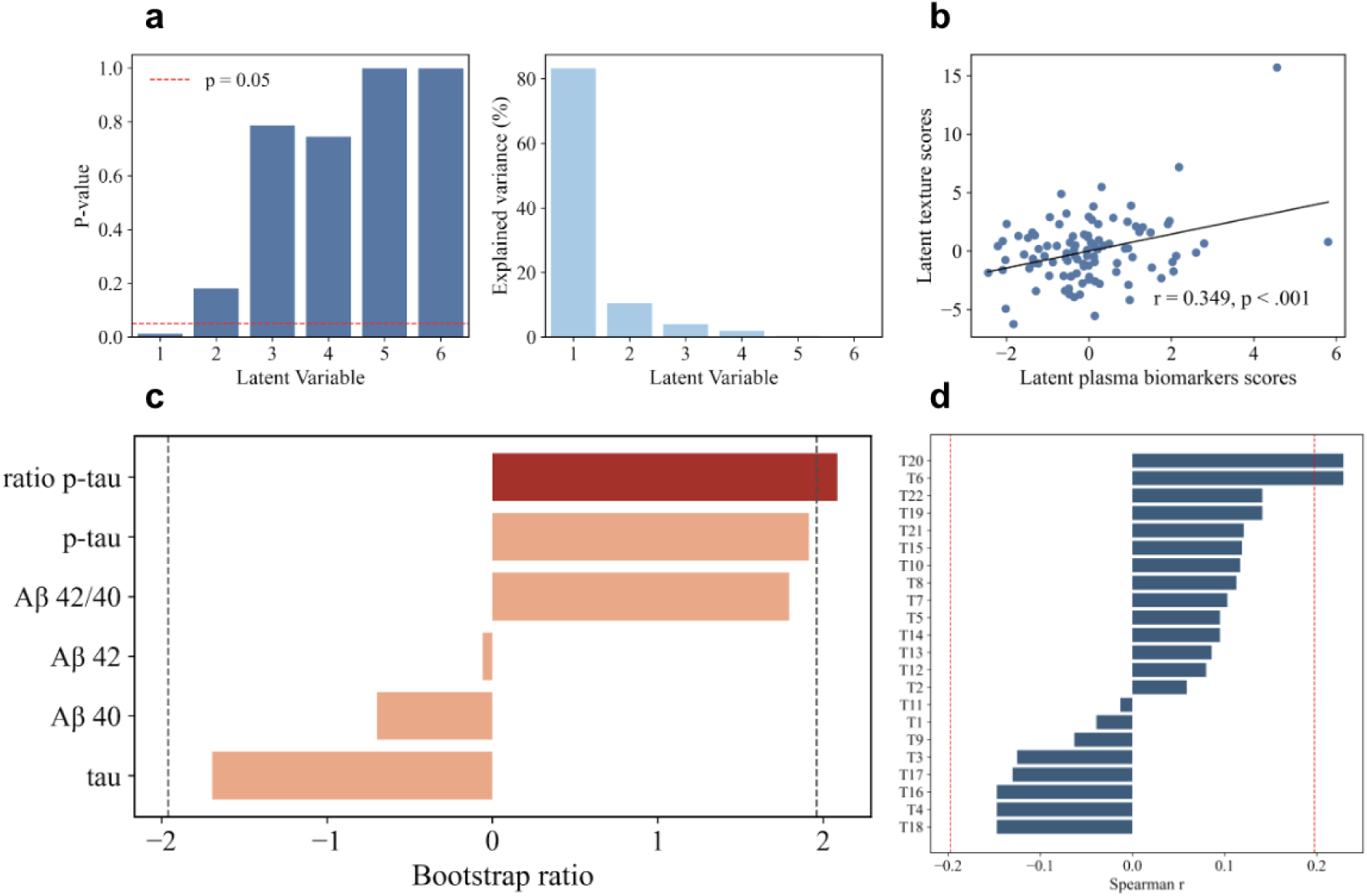
Partial least squares correlation (PLSC) results for latent variable 1 (LV1): Associations between training-related changes in left hippocampal textures and changes in plasma AD-related biomarkers. (a) P-values of latent variables, showing that only LV1 reached statistical significance (p < .05). (b) Correlation between latent texture scores and latent plasma biomarker scores. (c) Bootstrap ratios for plasma AD-related biomarker variables (X-side), reflecting the contributions of biomarker changes to LV1. (d) Spearman correlations between texture measures and plasma biomarker latent scores (Y-side). PLSC was conducted using post–pre difference scores for both texture and plasma biomarker variables. Red dashed lines in Panels B and D indicate the significance threshold (p = .05 or equivalent correlation cutoff). T1 = Uniformity/Energy/Angular Second Moment, T2 = Entropy, T3 = Dissimilarity, T4 = Contrast/Inertia, T5 = Inverse Difference, T6 = Correlation, T7 = Homogeneity/Inverse Difference Moment, T8 = Autocorrelation, T9 = Cluster Shade, T10 = Cluster Prominence, T11 = Maximum Probability, T12 = Sum of Squares, T13 = Sum Average, T14 = Sum Variance, T15 = Sum Entropy, T16 = Difference Variance, T17 = Difference Entropy, T18 = Information Measure of Correlation 1 (IMC1), T19 = Information Measure of Correlation 2 (IMC2), T20 = Maximal Correlation Coefficient, T21 = Inverse Difference Normalized (IDN), T22 = Inverse Difference Moment Normalized (IDMN).

On the texture side, hippocampal feature–latent score associations reflected a pattern of increasing microstructural regularity (Figure 8d). Correlation and maximal correlation coefficient showed positive associations with the latent brain score, whereas entropy-, variance-, and contrast-related measures were negatively associated, indicating reduced heterogeneity and enhanced tissue organization. Directionality was interpreted according to established feature semantics (Supplementary Table S1).

Collectively, these findings demonstrate that hippocampal microstructural reorganization following training—characterized by increasing regularity and decreasing disorder—is robustly associated with reductions in plasma p-tau–related biomarkers. This pattern suggests tight coupling between hippocampal structural plasticity and favorable molecular changes in tau-related pathology.

## 4. Discussion

The present study examined whether heart rate variability (HRV) biofeedback targeting increased heart rate oscillations (Osc+) is associated with changes in microstructural brain texture, an imaging marker shown to be sensitive to early Alzheimer’s disease (AD)–related alterations. Because texture was characterized using a broad set of 22 gray-level co-occurrence matrix (GLCM) features capturing partially overlapping aspects of tissue organization, we first evaluated intervention effects at the composite level using factor scores. This strategy allowed us to identify latent texture dimensions showing potential sensitivity to the intervention, after which we conducted follow-up analyses on individual texture indices contributing most strongly to the significant factors. Multivariate outliers were identified using a PCA-based approach. Statistical inference for ROI-based analyses was conducted using linear mixed-effects models while multivariate brain-plasma associations were assessed using partial least squares correlation.

Intervention effects on composite texture indices were selective and modest. In the ADSC, effects were confined to a single factor showing a borderline Time × Condition interaction, driven by opposing trajectories across Osc+ and Osc− conditions without significant within-group changes, and with only tentative evidence for age-related modulation. In contrast, the hippocampus showed a clearer age-dependent pattern, with a significant Time × Condition × Age Group interaction indicating that older adults, but not younger adults, exhibited condition-dependent changes in texture. Notably, these effects were expressed as relative differences between conditions rather than robust absolute changes, highlighting that HRV biofeedback influences microstructural texture in a subtle, interaction-driven manner.

We further examined the individual texture indices which revealed that the intervention effects were specifically expressed along a microstructural regularity dimension, captured by uniformity and entropy. In the ADSC, the combined pattern of increased uniformity and decreased entropy in the Osc+ condition suggests enhanced spatial organization and reduced heterogeneity of tissue signals, consistent with improved microstructural integrity. Although these effects were not robustly moderated by age at the model level, exploratory analyses indicated that they were largely driven by older adults, suggesting greater sensitivity to intervention in later life. This selective modulation of the uniformity–entropy dimension is consistent with prior work suggesting that microstructural texture features are particularly sensitive to early AD-related tissue alterations that are not fully captured by volumetric measures. In particular, increased tissue heterogeneity, reflected by lower uniformity and higher entropy, has been associated with early amyloid- and tau-related processes in both cortical and medial temporal regions (Cai et al., 2020b; Wearn et al., 2023).

In contrast, hippocampal effects were characterized by age-dependent and bidirectional changes, with significant three-way interactions indicating that intervention effects differed across age groups. In older adults, the opposing patterns of uniformity and entropy across conditions suggest that HRV manipulation may differentially influence hippocampal organization depending on the direction of training. However, because these effects were not supported by significant simple effects, they should be interpreted as relative condition differences rather than strong evidence of change within groups. Together, these findings indicate that HRV biofeedback may selectively modulate microstructural organization, but that its effects are regionally heterogeneous and more detectable in older adults.

Subregional analyses did not provide robust evidence for anatomically localized effects. Although several Time × Condition effects emerged at the uncorrected level, particularly in lateral temporal regions and the right hippocampus, none survived Benjamini–Hochberg correction, indicating that these findings may reflect false positives or unstable estimates. As such, the results do not support precise regional or hemispheric specificity, and instead suggest that the observed intervention effects are likely subtle and spatially distributed. These subregional patterns should therefore be considered exploratory and hypothesis-generating.

Beyond these regional texture effects, multivariate analyses revealed robust and coordinated associations between texture alterations and plasma AD-related biomarkers. Across the entorhinal cortex, fusiform gyrus, and hippocampal regions, significant latent variables indicated that increases in texture regularity (e.g., higher homogeneity, and lower entropy and variance) were associated with decreases in tau- and p-tau-related biomarkers. This pattern was highly consistent across regions, with tau-related measures emerging as the primary drivers of this plasma profile, suggesting that the observed brain-plasma associations are specifically linked to tau-related processes. These associations were stronger and more consistent than the univariate intervention effects. Intervention-related microstructural changes may be subtle at the regional level but become more detectable when examined as coordinated multivariate patterns.

Importantly, prior work has shown that texture metrics provide information about dementia risk that is at least partly independent of conventional volumetric measures (Sørensen et al., 2015). In this context, the observed coupling between intervention-related texture changes and tau-related biomarkers suggests that HRV biofeedback may influence microstructural properties that are closely linked to underlying disease-relevant processes, rather than merely reflecting secondary consequences of macroscopic structural change. Thus, texture measures in the present study function not only as descriptive indices of tissue organization but also as functionally informative markers reflecting coordinated brain–molecular dynamics.

By demonstrating selective, age-dependent, and regionally specific modulation of microstructural texture, this study extends previous work linking HRV to macroscopic brain measures such as cortical thickness and volume. Rather than inducing generalized structural change, HRV biofeedback appears to influence specific microstructural properties that are sensitive to early AD-related alterations, particularly within medial and lateral temporal regions. These findings highlight microstructural texture as a promising target for physiological interventions and underscore the importance of focusing on intervention-specific trajectories and biologically grounded outcome measures when evaluating the neural impact of autonomic training.

## Data Availability

The raw data used in this study are publicly available through OpenNeuro as part of the HRV-ER dataset (https://openneuro.org/datasets/ds003823). The derived texture-analysis data generated for this study, as well as the code used for texture analysis, are available from the corresponding author upon reasonable request. Code used for statistical analyses is also available from the corresponding author upon request.

