## Supplementary Table S1 for "Effects of a 5-week heart rate biofeedback randomized intervention on texture in the Alzheimer’s Disease signature cortical region"

*GLCM Texture Feature Interpretation and Directionality*

| **Feature No.** | **Feature name** | **Interpretation** | **Preferred direction** | **Biological meaning** |
| --- | --- | --- | --- | --- |
| T1 | Uniformity / Energy / Angular Second Moment | Global homogeneity, uniform gray-level distribution | Higher = better | More regular and homogeneous tissue organization |
| T2 | Entropy | Randomness / disorder | Lower = better | Less heterogeneous, more organized tissue |
| T3 | Dissimilarity | Local intensity variation | Lower = better | Smoother local texture, reduced microstructural irregularity |
| T4 | Contrast / Inertia | Edge strength, high-frequency variation | Lower = better | Reduced sharp transitions, greater structural smoothness |
| T5 | Inverse Difference | Local similarity | Higher = better | Greater similarity between neighboring voxels |
| T6 | Correlation | Linear dependency between neighboring voxels | Higher = better | Greater structural coherence |
| T7 | Homogeneity / Inverse Difference Moment | Weighting toward similar gray levels | Higher = better | More homogeneous tissue texture |
| T8 | Autocorrelation | Repetition of texture patterns | Higher = better | Greater regularity of spatial structure |
| T9 | Cluster Shade | Skewness / asymmetry of texture distribution | Lower = better | Reduced asymmetry, more stable distribution |
| T10 | Cluster Prominence | Peakedness / outlier sensitivity | Lower = better | Reduced extreme texture values |
| T11 | Maximum Probability | Dominant texture pattern | Higher = better | Stronger dominant homogeneous pattern |
| T12 | Sum of Squares (Variance) | Overall gray-level variability | Lower = better | Reduced intensity dispersion |
| T13 | Sum Average | Mean gray-level magnitude | Context dependent | Reflects average signal intensity; interpretation depends on modality |
| T14 | Sum Variance | Global variance around mean | Lower = better | Reduced heterogeneity |
| T15 | Sum Entropy | Global disorder | Lower = better | More ordered tissue structure |
| T16 | Difference Variance | Local variance of differences | Lower = better | Reduced microstructural variability |
| T17 | Difference Entropy | Local disorder | Lower = better | More organized local texture |
| T18 | Information Measure of Correlation (IMC1) | Information shared between voxels | Higher = better | Greater spatial dependency and structure |
| T19 | Information Measure of Correlation (IMC2) | Nonlinear voxel dependency | Higher = better | Stronger complex structural relationships |
| T20 | Maximal Correlation Coefficient | Maximal nonlinear correlation | Higher = better | Richer and more organized texture structure |
| T21 | Inverse Difference Normalized (IDN) | Normalized local homogeneity | Higher = better | Increased normalized similarity |
| T22 | Inverse Difference Moment Normalized (IDMN) | Normalized homogeneity measure | Higher = better | Greater normalized tissue regularity |

*Note.*  This table summarizes the biological interpretation and preferred directionality of gray-level co-occurrence matrix (GLCM) texture features used in the present study. Directionality reflects widely adopted conventions in neuroimaging and radiomics literature, in which higher values of features indexing homogeneity, similarity, and spatial dependency are generally interpreted as reflecting greater tissue regularity, whereas higher values of features indexing entropy, contrast, and variance are interpreted as reflecting greater disorder or heterogeneity (Haralick et al., 1973; Castellano et al., 2004; Thibault et al., 2014; Parekh & Jacobs, 2016). For context-dependent features, biological interpretation may vary depending on image contrast, acquisition parameters, and preprocessing.

**Supplementary Figure S1.**

*Flow Diagram of Randomized Trial*

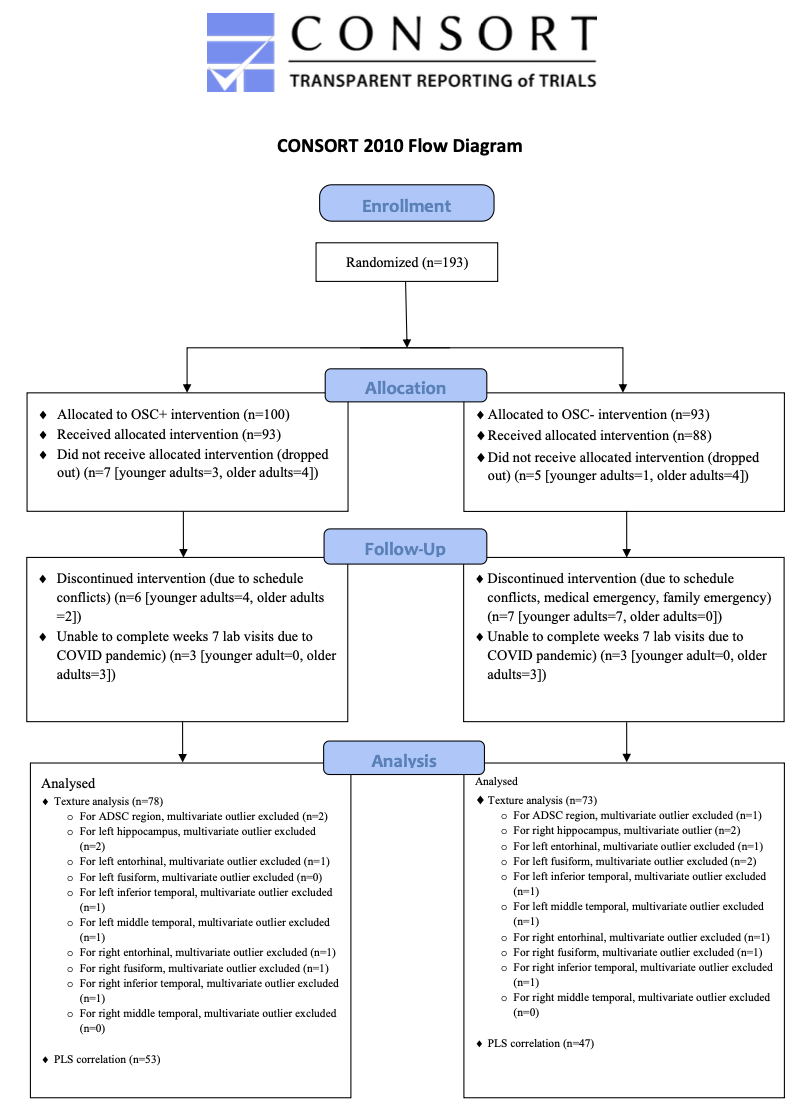
